# Symptomatic and asymptomatic transmission of SARS-CoV-2 in K-12 schools, British Columbia, April to June 2021

**DOI:** 10.1101/2021.11.15.21266284

**Authors:** Alex Choi, Louise C. Mâsse, Samantha Bardwell, Yanjie Zhao, Yang Xin Zi Xu, Ani Markarian, Daniel Coombs, Allison Watts, Adrienne Macdonald, Nalin Dhillon, Michael Irvine, Collette O’Reilly, Pascal M. Lavoie, David Goldfarb

## Abstract

We prospectively studied SARS-CoV-2 transmission at schools in an era of Variants of Concern (VoCs), offering all close contacts serial viral asymptomatic testing up to 14 days. Of 229 school close contacts, 3 tested positive (1.3%), of which 2 were detected through asymptomatic testing. Most secondary transmission (90%) occurred in households. Routine asymptomatic testing of close contacts should be examined in the context of local testing rates, preventive measures, programmatic costs, and health impacts of asymptomatic transmission.

---

Consistent with contact tracing studies done internationally, Canada has observed limited transmission of SARS-CoV-2 among children in school settings. (1–4) However, infection in children typically results in mild illness, and many cases may be minimally symptomatic or asymptomatic. (5,6) To assess asymptomatic transmission, a smaller number of studies have systematically tested students’ and staff members’ contacts. (7–9) While these studies have similarly detected minimal spread within schools, they have included a relatively low total number of index cases and were conducted before the emergence of more transmissible Variants of Concern (VoCs). Moreover, these studies did not account for differential risk between classmates, tested full classes instead of prioritizing contacts with higher levels of exposure (e.g. deskmates/close friends), and could underestimate the real, material risk by inflating the denominator.

The primary aim of this study was to assess the added yield of an asymptomatic serial testing program for close contacts of primary cases including those in the school setting.

## COVID-19 prevention measures in schools

Following closures in March 2020, K-12 schools in British Columbia reopened for the 2020/2021 academic year in September 2020. K-12 schools implemented COVID-19 safety plans developed with support from Public Health, which included public health measures, environmental measures, administrative measures, personal measures, and personal protective equipment (Appendix 1 and see details of prevention and control measures in published reports (1,4)). (10) Despite the emergence of Alpha, Beta, and Gamma VOCs, all schools remained open for the duration of the 2020/2021 school year.

## Identification of student and staff COVID-19 cases and their closest contacts

Positive or indeterminate Nucleic Acid Amplification Tests (NAAT) and VoC screening results for Vancouver students and staff were automatically reported to Public Health and evenly distributed to 2 contact tracing teams. This study included all lab-confirmed and probable-lab cases among students or staff and their contacts, assigned to contact tracing team 2 from April 12, 2021, until June 30, 2021. Case and contact definitions can be found in Appendix 2. Contact tracers obtained informed consent and collected case and contact information using a standardized form through telephone interviews of students/staff members and guardians, supplemented by collateral information from school districts, principals, and teachers

We longitudinally followed all close contacts, classroom contacts, and school contacts regardless of whether or not they underwent asymptomatic testing. For school exposures, individual risk assessments were conducted integrating cases’ symptoms, ages of cases and contacts, nature, and duration of contact, mask use, setting (e.g. indoor/outdoor), and presence/absence of known SARS-CoV-2 transmission. (11) Classmates specifically identified as close contacts by students, guardians or staff were asked to undergo free, asymptomatic SARS-CoV-2 NAAT, self-isolate for 14 days, and self-monitor for symptoms. This typically included those who were close to the case during the school day (e.g. deskmates) and close friends. (12) Asymptomatic testing was done using three serial, validated saline mouth rinse gargles: upon contact identification, 7-8 days after last exposure, and 10-14 days after last exposure. (13,14) All other classroom contacts were asked to self-monitor for symptoms. If symptoms emerged, classroom and close contacts were directed to seek immediate, additional testing through public clinics that provided free, rapidly processed, widely accessible symptomatic testing. (15) Entire classes were isolated and offered testing in the event of a school cluster where transmission may have occurred within the classroom.

To characterize clusters, primary cases were defined as laboratory-confirmed or lab-probable cases with the earliest symptom onset dates within the school setting (Appendix 2). Where a classmate of a primary case tested positive, linkage was assumed. Where contacts of classmates tested positive (e.g. family members of a classmate), repeat asymptomatic testing of the classmate was requested, and exposure through the primary student/staff case was assumed.

## Ethical statement

This project was reviewed by the BC Children’s and Women’s Research Ethics Board and deemed a QA/QI activity and was therefore exempted from formal ethical review but received a privacy impact assessment (#PIA 2021-40). Informed consent was obtained from all participants or their guardians.

## Transmission of SARS-CoV-2 from student and staff cases

No schools were closed during the study period. Characteristics of primary cases and their close contacts are described in Table 1. During the study period, 69 primary cases were identified among K-12 students and staff. Of these, 23 (33%) were female and 46 (67%) were male, 65 (94%) were students and four (6.2%) were staff. The majority of cases resided within Vancouver.

**Table 1.**
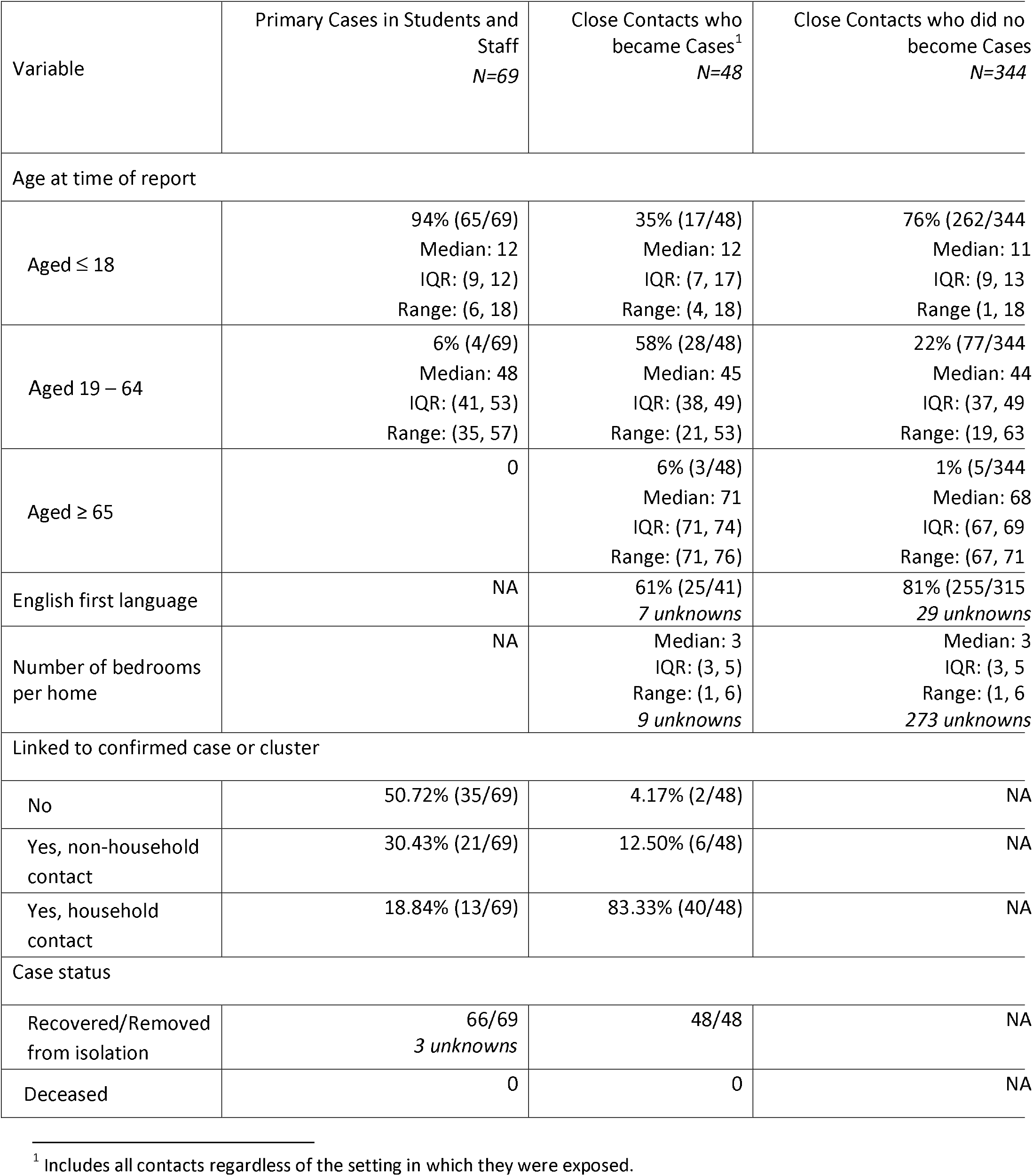
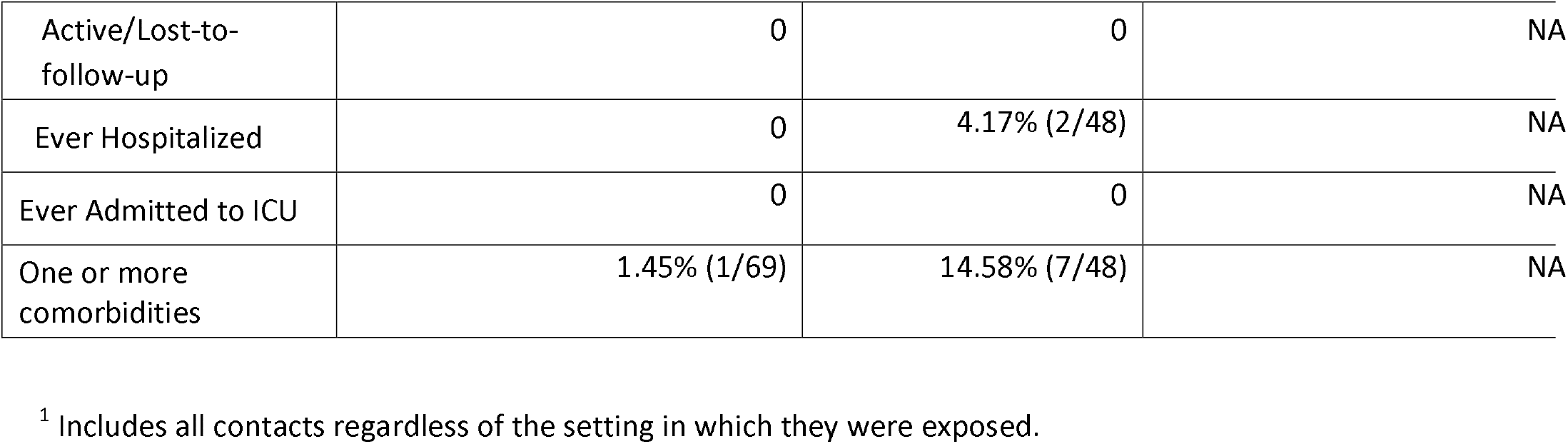
Characteristics of 69 student and staff SARS-CoV-2 cases and all close contacts.

**Table 2:** Characteristics of contact events between primary cases and school contacts.

|  | **School contacts who became**<br>**secondary cases**<br>**N = 3** | **School contacts who did not**<br>**become secondary cases**<br>**N = 226** |
| --- | --- | --- |
| **Details of contact** |  |  |
| Close proximity | 100% (3/3) | 67.70% (153/226) |
| Interacting | 100% (3/3) | 80.53% (182/226) |
| Touching | 33.33% (1/3) | 16.37% (37/226) |
| Secretions | 0% (0/3) | 1.77% (4/226) |
| **Any exercise or physical**<br>**activity with case** | 66.67% (2/3) | 27.88% (63/226) |
| **Duration of close contact** |  |  |
| Less than 15 minutes | 0% (0/3) | 1.87% (4/214) |
| 15 minutes to 1 hour | 0% (0/3) | 14.02% (30/214) |
| 1-2 hours | 33.33% (1/3) | 18.69% (40/214) |
| 2-4 hours | 0% (0/3) | 13.55% (29/214) |
| Over 4 hours | 66.67% (2/3) | 34.11% (73/214) |
| Unknown* | 0% (0/3) | 23.4% (50/214) |
| **Locations where contact**<br>**occurred** |  |  |
| Indoors | 100% (3/3) | 41.59% (94/226) |
| Outdoors | 100% (3/3) | 83.63% (189/226) |
| Motor vehicle/bus | 0% (0/3) | 1.33% (3/226) |
| **Primary case was wearing a**<br>**mask** |  |  |
| Not wearing a mask | 66.67% (2/3) | 17.13% (37/216) |
| Wearing a mask | 0% (0/3) | 40.74% (88/216) |
| Sometimes wearing a mask | 33.33% (1/3) | 38.89% (84/216) |
| Unknown* | 0% (0/3) | 7.87% (17/216) |

\* Where no response was indicated, “unknown” was used.

Based on the contact tracing of 69 primary cases, 392 close contacts were identified, offered asymptomatic testing, and instructed to self-isolate. Of these 392 close contacts, 229 were school contacts, 117 were household contacts, 22 were social contacts, 3 were extracurricular contacts, 16 contacts were mixed (school and social), and the nature of the close contact could not be identified in 5 cases (Figure 1). One hundred and sixty-eight close contacts (43%) participated in asymptomatic testing, and 224 (57%) declined asymptomatic testing but agreed to participate in data collection. Of the 168 who agreed to asymptomatic testing, 13 (7.7%) tested positive. Of these 13 secondary cases, 2 were school contacts (15%), 10 were household contacts (77%) and 1 was a mixed school and social contact (8%). During the study 224 declined asymptomatic testing, and 35 (16%) later tested positive through the public symptomatic testing system. Of 229 school contacts, 66.3% spent over 1 hour with the case, 83.6% interacted with the case indoors, and 56% wore a mask sometimes or not at all; 1.3% became secondary cases.

**Figure 1:**
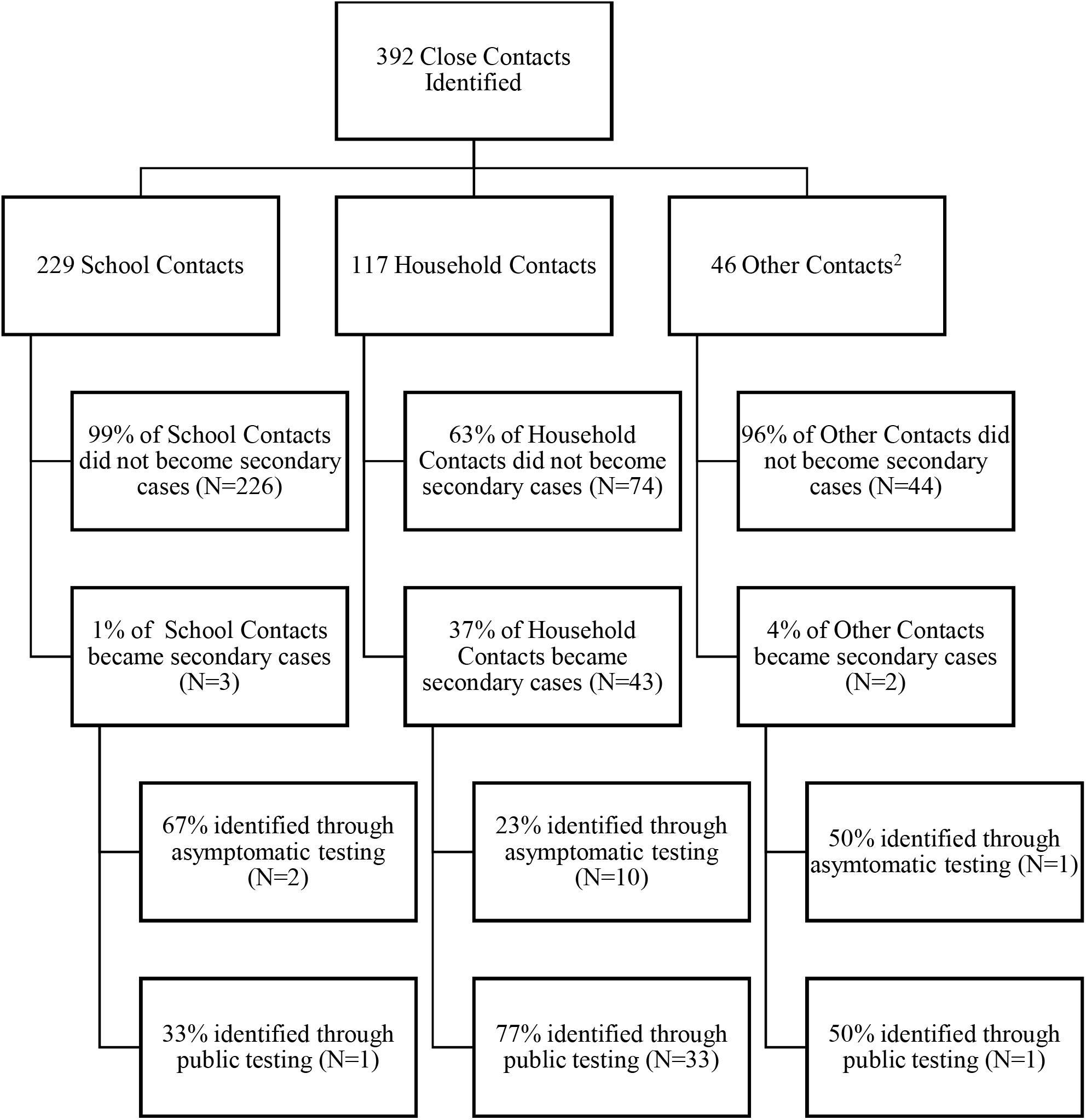
Flow diagram of close contacts who became cases and who did not by contact type.

Upon review of symptom data (Figure 2), 5 of the 13 (38%) cases identified through asymptomatic testing were pre-symptomatic, meaning that they showed no symptoms at the time of close contact identification, but developed symptoms later. Three (23%) were minimally symptomatic, describing discomfort or non-specific illness but no discrete symptoms. A further 3 (23%) cases described clear symptoms. Two (15%) cases remained asymptomatic during the entire follow-up period (10 days after testing positive).

**Figure 2:**
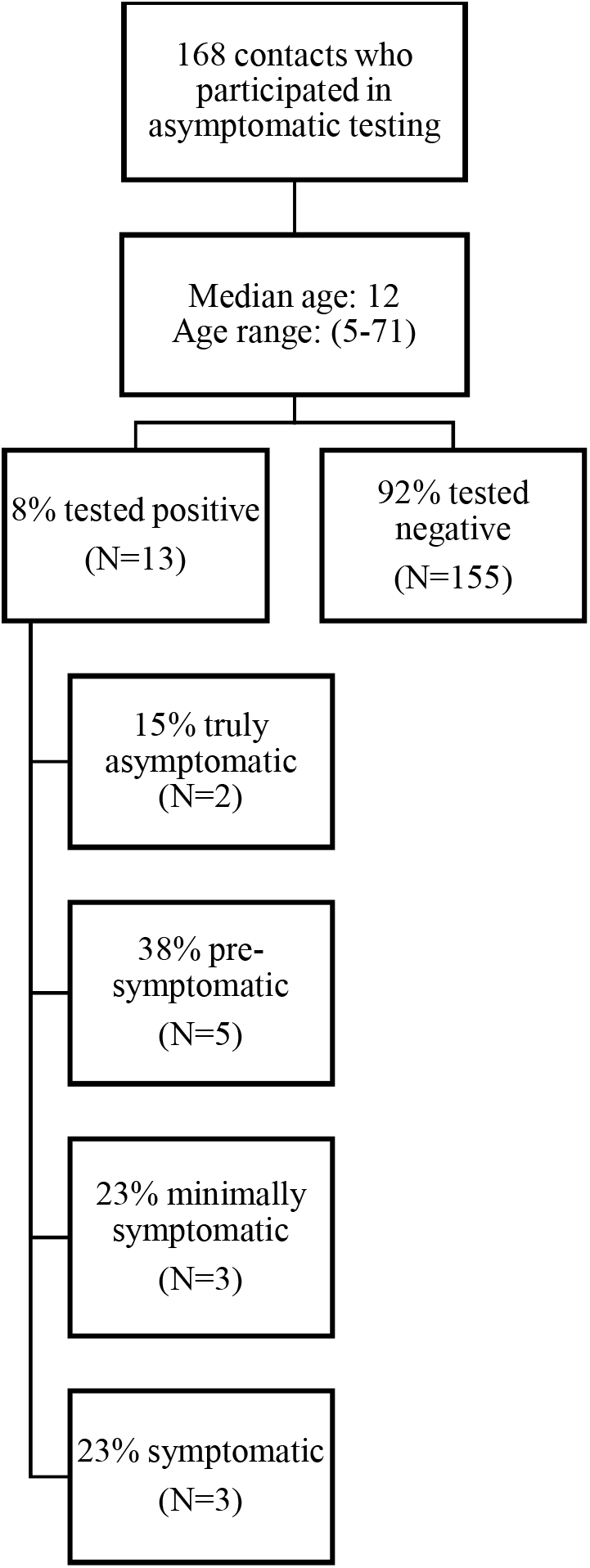
Flow chart of asymptomatic testing paritipation and test results.

Aggregating the asymptomatic and symptomatic testing results in 392 close contacts, 48 (12%) were identified as secondary cases. The majority (43/48 cases, 90%) were household contacts. In follow-up of 229 school contacts with the closest interactions with the primary case, 3 (1.3%) became secondary cases.

## Discussion

To our knowledge, our study is one of few that have systematically examined asymptomatic transmission in the context of VoCs. We found a low proportion of secondary cases among close contacts of primary cases identified in the school setting, even accounting for asymptomatic cases. Moreover, we found that the majority of the transmission occurred outside of school, and household contacts had the highest secondary transmission rate. Of note, the secondary transmission rate in schools in our study was higher than previously observed, which could be expected given that testing was limited to classmates with the closest interactions. (7,8) However, the yield of asymptomatic testing remained low even amongst classmates with prolonged close contact. Given that so few school contacts went on to become secondary cases, and the potential harms that can arise from missing school, this data questions the value of isolation as a measure for controlling SARS-CoV-2 transmission within the school setting. The low numbers of social and extracurricular contacts identified reflected limitations placed on social gatherings and extracurricular activities during the study period, and prevented risk comparisons.

A major strength of our study is that it includes data collection and prospective monitoring for a larger number of primary cases from both public and independent schools. However, our study is limited by the relatively low proportion (42.9%) of close contacts who agreed to undergo asymptomatic testing. Low participation may reflect operational challenges that could be more pronounced in communities with existing barriers to health care. Further research investigating barriers to testing and subpopulations that may derive more benefit from enhanced testing may be warranted.

## Conclusion

Enhanced and facilitated contact tracing and asymptomatic testing suggest low transmission of SARS-CoV-2 in K-12 schools with communicable disease prevention measures in the era of VoCs. However, two out of three school contacts identified as secondary cases were found through asymptomatic testing. Asymptomatic testing may be a useful adjunct, particularly where barriers to testing exist. Acknowledging resource requirements of asymptomatic testing including sufficient staffing, availability of test kits, and test processing capacity, the benefits of asymptomatic testing may need to be balanced against barriers to participation and costs.

## Data Availability

All data produced in the present study are available upon reasonable request to the authors.

## Acknowledgements

The authors would like to thank all of the school staff who distributed and completed the survey. The authors are also grateful to the school district and contact tracing staff who made this work possible: Kathy O’Sullivan, Tracy Au-Yeung, and the contact tracers and leads in pod 2. The study was funded by the Federal Government of Canada via its COVID-19 Immunity Task Force (CITF). PML and LCM receive salary support from the BC Children’s Hospital Foundation through Investigator Grant Award Programs.

