## Appendix 2 for "Symptomatic and asymptomatic transmission of SARS-CoV-2 in K-12 schools, British Columbia, April to June 2021"

Appendix 2. Data Dictionary

| **Term** | **Definition** |
| --- | --- |
| Lab confirmed case | A person with laboratory confirmation of infection with the virus that causes COVID-19 performed at a community, hospital or reference laboratory (NML or a provincial public health laboratory) running a validated assay. This consists of detection of at least one specific gene target by a NAAT assay (e.g. real-time PCR or nucleic acid sequencing).(45) |
| Probable-lab case | A person (who has had a laboratory test) with fever (over 38 degrees Celsius) or new onset of (or exacerbation of chronic) cough AND who meets the COVID-19 exposure criteria and in whom a laboratory diagnosis of COVID-19 is inconclusive. Inconclusive is defined as an indeterminate test on a single or multiple real-time PCR target(s) without sequencing confirmation or a positive test with an assay that has limited performance data available.(45) |
| Probable epi-linked case | A person (who has not had a laboratory test) with fever (over 38 degrees Celsius) or new onset of (or exacerbation of chronic) cough AND either close contact with a confirmed case of COVID-19 or lived in or worked in a closed facility known to be experiencing an outbreak of COVID-19 (e.g., long-term care facility, prison).(45) |
| Staff case | Case reporting working in a K-12 school within the learning setting in-person, or directly interacting with those within the learning setting, including teachers, support workers, principals/vice principals and office staff members. |
| Student case | Case (or proxy) reporting attending a K-12 school in-person. |
| Contact | Individual who had contact with a student or staff case during said case’s infectious period. From April 12, 2021 until May 8, 2021 only contacts who lived in the VCH region were eligible for inclusion. Beginning on May 9, contacts who attended or worked in schools in Vancouver were included regardless of their place of residence. Those who were already identified as a close contact of a case within the past two weeks were excluded unless they were part of a school cluster. |
| Primary case | Student or staff case with the earliest symptom onset date within the school setting. For asymptomatic cases, their test date was considered to be their symptom onset date. |
| Secondary case | Close contacts with SARS-CoV-2 infection detected through NAAT, whether symptomatic or asymptomatic. |
| Symptom onset date | Date when the patient’s first symptom emerged. For asymptomatic cases, their test date was considered to be their symptom onset date. |
| COVID-19 Symptoms | Case reported any one of: abdominal pain, arthralgia, chills, coma, confusion, conjunctivitis, cough, diarrhea, discoloration of toes or fingers, dizziness, fatigue, fever, headache, loss of appetite loss of sense of smell/taste, myalgia, nasal congestion, nausea, pharyngitis, rash, rhinorrhea, shortness of breath/breathing difficulty, or vomiting.(45) |
| Comorbidities | Case self-reported malignancy or cancer diagnosed in the last 5 years, chronic cardiac disease (excluding hypertension), diabetes, immunocompromised, pregnancy, chronic respiratory or pulmonary condition (excluding asthma) or smoking/vaping. |
| Lost to follow-up | Unable to complete follow up and confirm clinical status due to inability to contact patient. |
| Ever Hospitalized | Case was admitted to a hospital for at least an overnight stay, or with a prolongation of hospitalization, for reasons directly or indirectly related to their COVID-19 infection, and with no period of complete recovery between illness and admission.(45) |
| Ever Admitted to ICU | Case was admitted to an intensive care unit (ICU) for at least an overnight stay, or with a prolongation of ICU stay, for reasons directly or indirectly related to their COVID-19 infection, and with no period of complete recovery between illness and admission.(45) |
| Deceased | Death from any cause occurring in a person diagnosed with COVID-19 with no period of complete recovery between illness and death.(45) |
| Linked to confirmed case | Case reported close contact with a probable or confirmed case of COVID-19 within 14 days prior to illness onset. |
| Linked to confirmed case: School contact | Case (or proxy, teacher, or school administrator) reported close contact with a probable or confirmed case of COVID-19 in a school setting within 14 days prior to illness onset. This included all settings on and off school property that were used during the course of the school day, and included contact on school buses, in classrooms, at lunch/recess, and on field trips. It did not include extracurricular activities, which were recorded separately. |
| Linked to confirmed case: Social contact | Case (or proxy) reported close contact with a probable or confirmed case of COVID-19 in a social setting outside of the school within 14 days prior to illness onset. This included small and large group social events such as playdates, sleepovers, and parties, but did not include socialization during the school day (e.g. lunch/recess). |
| Linked to confirmed case: Household contact | Case reported close contact with a probable or confirmed case of COVID-19 in a household setting within 14 days prior to illness onset. |
| Linked to confirmed case: Extracurricular contact | Case (or proxy) reported close contact with a probable or confirmed case of COVID-19 in an extracurricular activity within 14 days prior to illness onset. Extracurricular activities are defined as extra academic or cultural activities performed by students outside of normal school curricula.This included extracurricular activities hosted by the school as well as those led by external organizations (e.g. sports teams). |
| Linked to confirmed case: Mixed contact | Case (or proxy) reported close contact with a probable or confirmed case of COVID-19 in two or more contexts – school, social, extracurricular, and household – within 14 days prior to illness onset. |
| Linked to confirmed case: No | Client did not report close contact with a probable or confirmed case of COVID-19 within 14 days prior to illness onset. |
| Cohort | A group of students and staff who remain together throughout a school term, the composition of which is consistent for all activities that occur in schools (40). While classes are the primary form of grouping where students spend the majority of their time, classes within a shared cohort may interact with each other for activities like physical education or music. |
| Close proximity | School contact was within 2 metres of primary student or staff case during their infectious period as per contact tracing interview. |
| Interacting | School contact was engaged in an action that involved the primary student or staff case during their infectious period as per the contact tracing interview. This could include talking, working, or playing. |
| Touching | School contact was directly touching primary student or staff case during their infectious period as per contact tracing interview. |
| Secretions | School contact was exposed to the respiratory secretions of the primary student or staff case during their infectious period as per contact tracing interview. This could include kissing, sharing cigarettes/vapes, or being coughed/sneezed upon. |
| Any exercise or physical activity with the case | School contact and primary student or staff case jointly engaged in activities requiring physical effort together (e.g. sports) during the infectious period of the student or staff case. |
| Duration of close contact | Period of time during which the school contact and the primary student or staff case were at a distance of less than 2 metres. |
