## Appendix 1a for "Symptomatic and asymptomatic transmission of SARS-CoV-2 in K-12 schools, British Columbia, April to June 2021"

##### COVID-19 Public Health Guidance for K-12 Schools

UPDATED: February 4, 2021

### Executive Summary

This document provides guidance for educators, administrators and support staff (hereafter referred to as staff) at public, independent and First Nations Kindergarten to Grade 12 (K-12) schools to minimize the transmission of COVID-19 and maintain a safe and healthy school environment for students, families and staff.

Our experience to date within BC, as well as international evidence, suggests that schools continue to be low-risk sites for COVID-19 transmission, even with increased risk of COVID-19 in some communities. However, while COVID-19 is present in our communities, it will exist in some schools.

The infection prevention and exposure control measures in place have been shown to be effective at stopping or limiting transmission of COVID-19 within schools. However, there are areas where renewed attention and focus are needed. These include:

1. Prevent crowding at all times; pay particular attention at the start and end of day.
2. Avoid close face-to-face contact whenever possible.
3. Assign staff to a specific cohort whenever possible.
4. Stagger recess, lunch and class transition times whenever possible.
5. Ensure that the use of masks does not reduce or replace practicing physical distancing and other prevention measures, for both students and staff.
6. Ensure prevention measures are in place in staff-only areas, including break and meeting rooms.
7. Implement music classes according to the [British Columbia Music Educators’ Association and the Coalition for Music Education in British Columbia](https://drive.google.com/file/d/1KG2rE1rU-NENxbQsuYN20xnM9TBlNn3Z/view) Guidance for Music Classes.
8. Ensure physical activity is delivered in line with the guidance in this document.

The Ministry of Education worked with Indigenous rights holders and K-12 education and health partners to build on public health guidance to establish the [Provincial COVID-19 Health and Safety Guidelines for K-12 School Settings](https://www2.gov.bc.ca/assets/gov/education/administration/kindergarten-to-grade-12/safe-caring-orderly/k-12-covid-19-health-safety-guidlines.pdf). These guidelines must be followed by schools and school districts, including if there are any differences between them and this public health guidance.

[WorkSafe BC guidance for offices](https://www.worksafebc.com/en/about-us/covid-19-updates/covid-19-returning-safe-operation/offices) lists measures that should be considered and implemented as applicable to the workplace for staff in office environments (both inside and outside of school buildings).

Contents

COVID-19 Public Health Guidance for K-12 Schools	1

Executive Summary	1

Introduction	4

Updates Within This Document	5

Supportive School Environments	5

Infection Prevention and Exposure Control Measures	6

Public Health Measures	8

Gatherings and Events	8

Public Health Case Management	8

Environmental Measures	10

Ventilation and Air Exchange	10

Space Arrangement	10

Cleaning and Disinfection	10

Traffic Flow	11

Physical Barriers	11

Administrative Measures	11

Cohorts	11

Physical Distancing	13

Staff-Specific Considerations	14

Other Prevention Strategies	14

Visitors	15

Curriculum, Programs and Activities	15

Student Transportation on School Buses	16

Food Services	17

Community Use of Schools	18

Personal Measures	18

Self Isolation and Illness	18

Hand Hygiene	20

Respiratory Etiquette	21

Water Stations and Fountains	21

Personal Items and School Supplies	21

Personal Protective Equipment (PPE)	21

Non-Medical Masks and Face Coverings (Masks)	21

PPE When Providing Student Services	22

Additional PPE	23

Appendix A: Evidence Summary	24

Appendix B: COVID-19 School Health and Safety Checklist	27

Appendix C: Contact Tracing and Public Health Notifications in Schools	30

Appendix D: Supplementary Guidance for School Meal Programs	31

Appendix E: What to Do if a Student or Staff Member Develops Symptoms At School	32

Appendix F: When to Perform Hand Hygiene at School	33

### Introduction

This document provides guidance for educators, administrators and support staff (hereafter referred to as staff) at public, independent and First Nations Kindergarten to Grade 12 (K-12) schools to minimize the transmission of COVID-19 and maintain a safe and healthy school environment for students, families and staff.

The Ministry of Education worked with Indigenous rights holders and K-12 education and health partners to build on public health guidance to establish the [Provincial COVID-19 Health and Safety Guidelines for K-12 School Settings](https://www2.gov.bc.ca/assets/gov/education/administration/kindergarten-to-grade-12/safe-caring-orderly/k-12-covid-19-health-safety-guidlines.pdf). These guidelines must be followed by schools and school districts, including if there are any differences between them and this public health guidance.

Supporting students to receive full-time, in-person learning offers considerable societal and individual benefits, particularly for those who already experience social and educational inequities. The benefits need to be balanced against the potential risk of COVID-19 spread and any evidence of benefit from school closures. Additional information is available from the BCCDC September 2020 report [Impact of School Closures on Learning, Child and Family Well-Being During the COVID-19 Pandemic](http://www.bccdc.ca/Health-Info-Site/Documents/Public_health_COVID-19_reports/Impact_School_Closures_COVID-19.pdf). Ensuring implementation and adherence to health and safety plans is important to keep in-person learning available.

During the fall, experience accumulated from around the world about the importance of keeping schools open and how to do so safely. Our experience to date within [BC](http://www.bccdc.ca/health-info/diseases-conditions/covid-19/data), [Canada](https://www.nccmt.ca/covid-19/covid-19-rapid-evidence-service/19), in [Europe](https://www.ecdc.europa.eu/en/publications-data/children-and-school-settings-covid-19-transmission) and [internationally](https://www.who.int/publications/m/item/weekly-epidemiological-update---19-january-2021), shows that schools continue to be low-risk sites for COVID-19 transmission, even with increased risk of COVID-19 in some communities. Since schools, childcare and other workplaces re-opened in the fall with comprehensive safety plans in place, we have seen that these settings carry a lower risk of transmission of COVID-19 compared to other settings where appropriate safety plans are not in place or are not consistently implemented. While COVID-19 is present in our communities, there will be students and staff with COVID-19 in schools. Local public health officials (school medical health officers) consistently monitor cases of COVID-19 in schools and support school communities to manage cases if and when they occur. A summary of current evidence is included as Appendix A.

School medical health officers noted that most school exposures in the fall did not result in transmission within the school. However, some school exposures did result in additional cases, including clusters and a small number of outbreaks. While each case was unique, school medical health officers recommend a review of existing health and safety measures to ensure all recommended measures are consistently implemented. It is recommended that all schools review their health and safety plans, with a focus on areas where measures may be absent or inconsistently implemented. The School Health and Safety Checklist included as Appendix B can be used to support these efforts. School medical health officers recommend the following as key areas of focus:

1. Prevent crowding at all times; pay particular attention at the start and end of day.
2. Avoid close face-to-face contact whenever possible.
3. Assign staff to a specific cohort whenever possible.
4. Stagger recess, lunch and class-transition times whenever possible.
5. Ensure that the use of masks does not reduce or replace practicing physical distancing and other prevention measures, for both students and staff.
6. Ensure prevention measures are in place in staff only areas, including break and meeting rooms.
7. Implement music classes according to the [British Columbia Music Educators’ Association and the Coalition for Music Education in British Columbia](https://drive.google.com/file/d/1KG2rE1rU-NENxbQsuYN20xnM9TBlNn3Z/view) Guidance for Music Classes.
8. Ensure physical activity is delivered in line with the guidance in this document.

The guidance included in this document should be implemented at all times within the school.

#### *Updates Within This Document*

This document is an update to guidance published on September 11, 2020. Changes are highlighted in yellow.

This document uses the terms elementary, middle and secondary to identify different approaches (where relevant) for schools based on the age range of students within them and the way learning is typically structured. If a school is unsure of which guidance to follow or these distinctions aren’t suitable to their school community, they can connect with their school medical health officer to determine what approaches are most suitable.

The term ‘mask’ in this document means a non-medical mask or face covering. Medical-grade masks are not recommended within school settings for general use.

The guidance in this document may not be relevant to distributed learning (including learning in non-traditional settings) or homeschooling. Administrators and leaders for those settings are encouraged to use guidance from this document, as well as guidance issued for other sectors as relevant, to reduce the risk of COVID-19 transmission in their unique environments.

BCCDC is the primary source of information about COVID-19 in BC. Resources on the BCCDC website can be used to support learning and to respond to questions you may receive from members of your school community. More information is available [here](http://www.bccdc.ca/health-info/diseases-conditions/covid-19/childcare-schools).

### Supportive School Environments

Schools can support students to practice personal preventive measures like physical distancing, hand hygiene, respiratory etiquette and mask use by:

- Having staff model these behaviours.
- Sharing reliable information to parents, families and caregivers. Information and resources are available from [BCCDC](http://www.bccdc.ca/health-info/diseases-conditions/covid-19/).
- Promoting them in the school through the use of visual aids like floor markings and signage.

Schools are encouraged to support student personal practices using positive and inclusive approaches. Schools should avoid punitive measures or enforcement activities that exclude students from fully participating in school or that could result in stigma.

### Infection Prevention and Exposure Control Measures

Infection prevention and exposure control measures help create safe environments by reducing the spread of communicable diseases like COVID-19. These are more effective in controlled environments where multiple measures of various effectiveness can be routinely and consistently implemented.

Schools are considered controlled environments. This is because schools include a consistent grouping of people, there are robust illness policies for sick students and staff and there is an ability to implement effective personal practices that are followed by most people most of the time in the setting (e.g. diligent hand hygiene, respiratory etiquette, etc.). This makes them different from public places like retail stores and public transit.

*The Hierarchy for Infection Prevention and Exposure Control Measures for Communicable Disease* describes measures to reduce the transmission of COVID-19 in schools. Control measures at the top are more effective and protective than those at the bottom. By implementing a combination of measures at each level, the risk of COVID-19 is substantially reduced.

**The Hierarchy for Infection Prevention and Exposure Control Measures for Communicable Disease**

**Public Health Measures**

**Includes orders from the Provincial Health Officer, improved testing, and contact tracing.**

**Environmental Measures**

**Includes being outdoors, physical barriers, visual cues for traffic flow and more frequent cleaning and disinfection.**

**Administrative Measures**

**Includes changes in scheduling and work practices, health and wellness policies, and placing students and staff in cohorts.**

**Personal Measures**

**Includes staying home when sick, and practicing physical distancing, hand hygiene and respiratory etiquette.**

**Personal Protective Equipment**

**Includes gloves and masks.**

**Less Effective	More Effective**

**Public Health Measures** are actions taken across society at the population level to limit the spread and reduce the impact of COVID-19. Public health has implemented broad public health measures, including: prohibiting large gatherings and events, restricting gatherings in homes, requiring travellers returning from outside of Canada to self-isolate or quarantine upon arrival in BC, effective case finding and contact tracing, and advising people to stay home when they are sick. Under the direction of Medical Health Officers, effective case finding and contact tracing is in place and prepared to manage any cases and confirmed contacts in the school setting.

**Environmental Measures** are changes to the physical environment that reduce the risk of exposure. Examples include being in outdoor spaces, ensuring good ventilation and air exchange, using visual cues for physical distancing or directing traffic flow in hallways, erecting physical barriers where appropriate and frequent cleaning and disinfection.

**Administrative Measures** are measures enabled through the implementation of policies, procedures, training and education that reduce the risk of exposure. Examples of these include health and wellness policies, working or learning in defined groups (cohorts), modified schedules and supporting the ability of individuals to practice physical distancing.

**Personal Measures** are actions individuals can take to protect themselves and others. Examples include practicing physical distancing, washing hands frequently, coughing into elbows and staying home if sick.

**Personal Protective Equipment** (PPE) can reduce the risk of COVID-19 transmission; however, it is not sufficient as a stand-alone preventive measure. It should be suited to the task, and must be worn and disposed of properly.

***Schools can implement a combination of measures at different levels, as described in this document. This document includes Environmental, Administrative, Personal Measures and the use of PPE.***

### Public Health Measures

#### *Gatherings and Events*

The [Provincial Health Officer’s Order for Gatherings and Events](https://www2.gov.bc.ca/assets/gov/health/about-bc-s-health-care-system/office-of-the-provincial-health-officer/covid-19/covid-19-pho-order-gatherings-events.pdf) is focused on one-time or recurrent events where people gather and where control measures may be hard to implement. It is updated as needed to reflect the management of COVID-19 in BC. This order does not apply to students, teachers or instructors at a school operating under the *School Act* [RSBC 1996] Ch. 412 or the *Independent School Act* [RSBC 1996] Ch. 216 or a First Nations School when engaged in educational activities. Educational activities include extracurricular activities offered by a school, but not social activities or community events held at a school.

###### School Gatherings

School gatherings are events that bring staff and students together outside of regular learning activities. School gatherings should occur within the cohort, and occur infrequently.

- Schools should seek virtual alternatives wherever possible when a group is gathered, including for assemblies, extracurricular practices, and other activities.
  - If a virtual alternative is not possible, the size of the gatherings should be minimized as much as possible.
  - Limit attendees from outside of the cohort to the minimum number necessary (e.g. school staff, visitors etc.) to meet its purpose.
- Examinations or assessments are not considered school gatherings. They should be implemented in accordance with the guidance for within-cohort and multi-cohort learning in this document.
- Gatherings or events at a school, including social gatherings of students and/or staff, must comply with the [PHO Order for Gatherings and Events](https://www2.gov.bc.ca/assets/gov/health/about-bc-s-health-care-system/office-of-the-provincial-health-officer/covid-19/covid-19-pho-order-gatherings-events.pdf).

#### *Public Health Case Management*

Public health teams actively monitor and trace cases of COVID-19 in schools.

###### Case Finding and Contact Tracing

If a staff or student is a confirmed case of COVID-19 through testing or investigation (i.e. case finding), public health will determine who that person has been in close contact with recently (i.e. contact tracing) to determine how they were infected and who else may have be at risk of infection.

###### Exposures and Exposure Notifications

An **exposure** occurs when a person attends school when they may have been potentially infectious with COVID-19 and there is a risk of transmission to others. If there was a potential exposure at a school, public health will work with the school to understand who may have been exposed, and to determine what actions should be taken, including identifying if other students or staff are sick (case finding) or have been exposed.

Public health will notify by phone or letter everyone who they determine may have been exposed, including if any follow-up actions are recommended. Those who public health determines are [close contacts](http://www.bccdc.ca/health-info/diseases-conditions/covid-19/self-isolation/close-contacts) will be advised to self-isolate. Those who public health determines are not close contacts but may still have been exposed may be advised to self-monitor for symptoms.

Once those who may have been exposed have been directly notified, regional health authorities post a notification on their website that details the school and dates a person attended when they may have been infectious. In some regions, this exposure notification is also provided by letter to the school community; this is determined at a regional level.

A process map for how contact tracing and public health notifications occur in schools is included as Appendix C.

To ensure personal privacy rights are maintained, public health will not disclose that a student or staff member is a confirmed case of COVID-19 unless there is reason to believe they may have been infectious when they attended school. Public health will only provide the personal information needed to support effective contact tracing.

School administrators or staff should not provide notifications to staff or students’ families about potential or confirmed COVID-19 cases unless the school administrator is directed to do so by the school medical health officer. School Administrators are to follow processes outlined in [COVID-19 Protocols for School and District Administrators: Management of Illness and Confirmed Cases.](https://www2.gov.bc.ca/assets/gov/education/administration/kindergarten-to-grade-12/safe-caring-orderly/covid-19-protocols-for-administrators.pdf)

###### Clusters

A **cluster** refers to two or more confirmed cases of COVID-19 that occur among students and/or staff within a 14-day period, and isolated transmission is suspected or confirmed to have occurred within the school. When this occurs, public health, under the direction of a Medical Health Officer will investigate to determine if additional measures are required to prevent transmission of COVID-19. It is expected that multiple cases may occur within a 14-day period, especially when COVID-19 is circulating in the community. This does not necessarily mean that transmission occurred in the school, as they can also be the result of interactions outside of the school setting.

###### COVID-19 Outbreaks in Schools

An **outbreak** is when there is sustained, uncontrolled, widespread transmission of COVID-19 within a school, and a Medical Health Officer determines extraordinary public health measures are necessary to stop further transmission in the school or school community. Extraordinary public health measures are at the discretion of the Medical Health Officer and may include ordering the school to close for a certain amount of time or requiring testing of all potentially exposed individuals regardless of symptoms.

###### School Health and Safety Checklist

Schools can use the School Health and Safety Checklist Tool included as Appendix B to ensure implementation of recommended measures.

###### Self-isolation and Quarantine

Information on self-isolation and quarantine, including for international travelers returning to BC, is available from [BCCDC](http://www.bccdc.ca/health-info/diseases-conditions/covid-19/self-isolation/contact-tracing).

###### Regional Differences

Specific regional requirements may be put in place by local Medical Health Officers to reflect differences in community transmission, based on their authority under provincial legislation.

### Environmental Measures

#### *Ventilation and Air Exchange*

At this time, there is no evidence that a building’s ventilation system, in good operating condition, would contribute to the spread of COVID-19.

Good indoor ventilation alone cannot protect people from exposure to the virus; however, it may reduce risk when used in addition to other preventive measures. For activities that take place indoors, application of the basic principles of good indoor air quality should continue, including supplying outdoor air to replenish indoor air by removing and diluting contaminants that naturally occur in indoor settings. All mechanical heating, ventilation and air conditioning (HVAC) systems should be checked to ensure they are working properly. Where possible, schools can open windows if weather permits and it doesn’t impact the functioning of ventilation systems.

For more information, see WorkSafe BC guidance on [general ventilation and air circulation](https://www.worksafebc.com/en/resources/about-us/covid-19/general-ventilation-and-air-circulation-covid-19-faq?lang=en).

#### *Space Arrangement*

Spread people out as much as possible. Consider different common space, classroom and learning environment configurations to allow space between students and adults (e.g., different desk and table formations).

Arrange desks/tables to maximize space between students. Seating arrangements where students directly face one another should be avoided, particularly for middle and secondary schools. Use consistent seating arrangements where possible.

Avoid activities that require close face-to-face contact during school activities.

#### *Cleaning and Disinfection*

Regular cleaning and disinfection are important to prevent the transmission of COVID-19 from contaminated objects and surfaces. Schools should be cleaned and disinfected in accordance with the BCCDC’s [Cleaning and Disinfectants for Public Settings](http://www.bccdc.ca/Health-Info-Site/Documents/CleaningDisinfecting_PublicSettings.pdf) document.

This includes:

- General cleaning and disinfecting of the premises at least **once every 24 hours.**
  - This includes items that only a single student uses, like an individual desk or locker.
- **Frequently-touched** surfaces should be cleaned and disinfected an additional time every 24 hours (i.e. twice total). At least one of these cleanings should occur during the school day.
  - These include door knobs, light switches, water fountains, toilet handles, tables, desks and chairs, keyboards, sports equipment, manipulatives and toys used by multiple students.
- Clean and disinfect any surface that is visibly dirty.
- Use common, commercially-available detergents and disinfectant products and closely follow the instructions on the label.
  - See Health Canada’s list of [hard-surface disinfectants for use against coronavirus (COVID-19)](https://www.canada.ca/en/health-canada/services/drugs-health-products/disinfectants/covid-19/list.html) for specific brands and disinfectant products.
- Limit frequently touched items that are not easily cleaned to those that support learning, health and development.
  - Some frequently touched items like toys or manipulatives may not be able to be cleaned often (e.g. fabrics) or at all (e.g. sand, foam, playdough, etc.). These items can continue to be used, if hand hygiene is practiced before and after use.
  - There is no evidence that the COVID-19 virus is transmitted via textbooks, paper or other paper-based products. Laminated or glossy paper-based products (e.g. children’s books or magazines) and items with plastic covers (e.g. DVDs) can be contaminated if handled by a person with COVID-19; however, the risk is low. There is no need for these items to be cleaned and disinfected or quarantined for any period of time, or for hand hygiene to be practiced before or after use.
- Additional guidance on cleaning electronics, toys, fabrics and other items not addressed in the Cleaning and Disinfectants for Public Settings resource is available [here](http://www.bccdc.ca/health-info/diseases-conditions/covid-19/prevention-risks/cleaning-and-disinfecting).
- Empty garbage containers daily and when full.
- Wear disposable gloves when cleaning blood or body fluids (e.g., runny nose, vomit, stool, urine). Wash hands before wearing and after removing gloves.

There are no additional cleaning and disinfecting procedures necessary. This includes when different cohorts use the same space (e.g. a classroom, gym, arts room, home economics or science lab, etc.).

#### *Traffic Flow*

Use floor markings and posters to direct traffic flow throughout the school. This may include one-way hallways and designated entrance and exit doors. It is important not to reduce the number of exits and to adhere to the fire code.

#### *Physical Barriers*

Barriers can be installed in places where physical distancing cannot regularly be practiced and a person is interacting with numerous individuals outside of a cohort. This may include the front reception desk where visitors check in or in the cafeteria where food is distributed. It may also include itinerant staff working across cohorts.

### Administrative Measures

Lowering the number of close, prolonged face-to-face interactions an individual has with different people helps to prevent the spread of COVID-19. This can be accomplished in K-12 school settings through two different but complementary approaches: cohorts (to reduce the number of potential interactions) and physical distancing (to change the nature of interactions).

#### *Cohorts*

A **cohort** is a group of students and staff who remain together throughout a school term. The use of cohorts in schools allows for a significant reduction in the number of individual interactions, while allowing most students to receive in-person learning in a close-to-normal school environment. Interactions within the cohort will vary, with classes continuing as the primary form of grouping where students will spend the majority of their time.

- In elementary and middle schools, a cohort can be composed of up to 60 people per quarter, semester or term.
- In secondary schools, a cohort can be composed of up to 120 people per quarter, semester or term.
- Cohorts can be composed of students and staff.

School administrators should determine the composition of the cohorts. The composition of the cohort should remain consistent for all activities that occur in schools, including but not limited to learning and breaks (lunch, recess, classroom changes, etc) during the quarter, semester or term.

Students with disabilities and diverse abilities may require unique considerations to ensure their inclusion in a cohort. Schools can adapt the guidance in this document as necessary to ensure the inclusion of these students while ensuring the intent is maintained. Schools can connect with their school medical health officer for support and guidance.

Cohort composition can be changed at the start of a new quarter, semester or term in the school year. Outside of these, composition should be changed as minimally as possible, except where needed to support optimal school functioning. This may include learning, operational or student health and safety considerations.

Consistent seating arrangements are encouraged within cohorts where practical.

School administrators should keep up-to-date lists of all members of a cohort and their contact information to support swift communications from the school and to share with public health to support contact tracing, if needed.

###### Multi-Cohort Learning

Multiple groups of students from different cohorts can be in the same learning space at the same time if physical distancing can be strictly practiced between people from different cohorts, and there is adequate space available to prevent crowding of those from within the same cohort.

Masks are not a replacement for physical distancing.

###### Multi-Cohort Services

Students from different cohorts may need to be together to receive beneficial social supports, programs or services (e.g. meal programs, after school clubs, etc.). Within these supports or services, it is expected that cohorts are maintained, and physical distancing is practiced as much as is practical to do so while still ensuring the support, program or service continues. This does not apply to extracurricular activities where physical distancing between cohorts should consistently be practiced. Guidance for when masks should be worn is available in the Personal Protective Equipment section of this document.

###### Outside of Cohort Social Interactions in Common Areas

Students and staff should do the following to safely socialize with those in different cohorts in common areas during transition times or break periods:

- In elementary schools, students can socialize with peers in different cohorts if they are outdoors and can minimize physical contact or if they are indoors and can practice physical distancing.
  - Elementary students are less able to consistently practice physical distancing. Outdoors is a lower-risk environment than indoors.
- In middle and secondary schools, students can socialize with peers in different cohorts if they can practice physical distancing.
  - Middle and secondary school students are expected to be capable of practicing physical distancing. If a student is unable to do so, they should socialize within their cohort or where they can be supported to practice physical distancing.
- Staff at all schools should seek to reduce the number of close, face-to-face interactions with each other at all times, even if wearing a mask or working within the same cohort. This includes during social interactions in staff areas and during meetings.

#### *Physical Distancing*

**Physical distancing** refers to a range of measures aimed at reducing close contact with others. Physical distancing is used as a prevention measure because COVID-19 tends to spread through prolonged, face-to-face contact.

- Within cohorts, physical distancing should include avoiding physical contact, minimizing close, prolonged, face-to-face interactions, and spreading out as much as possible within the space available.
  - Young children may not be able to consistently reduce physical contact.
- Outside of cohorts, practicing physical distancing should include avoiding physical contact and close, prolonged face-to-face interactions, spreading out as much as possible within the space available, and ensuring there is 2 meters of space available between people.
- For situations where members of different cohorts interact:
  - If people will be in the same space for an extended period of time (e.g. beyond 15 minutes), the space should be sufficiently large, and/or should have limits on the number of people so that 2 meters of space is available between people from different cohorts.
  - If people will be in the same space for transition purposes (e.g. changing between classes), and other measures are in place (e.g. markings on the floor, staggered transition times), there should be enough space to ensure no physical contact.

Elementary and middle schools are likely able to implement cohorts without reducing the number of individuals typically within the school. Secondary schools may use both approaches: implement cohorts and reduce the number of individuals typically within the school to ensure there is space available to prevent crowding. This may be necessary due to the larger number of people and the increased frequency of classroom exchanges that typically occur within secondary schools. Secondary schools should continue to prioritize the attendance of students who most benefit from in-person support and learners with diverse needs, as well as consider alternative learning modalities and off-campus learning.

Masks are not a replacement for physical distancing. Efforts should continue to focus on using all available space and preventing crowding or close gatherings.

Guidance for when masks should be worn is available in the Personal Protective Equipment section of this document.

#### Staff-Specific Considerations

###### Itinerant Staff

Schools should seek to assign staff to a single cohort whenever possible. This is intended to minimize the number of adults (staff and others) who interact with cohorts of which they are not a part.

Staff not assigned to a single cohort should practice physical distancing when interacting with each cohort. If physical distancing cannot consistently be practiced when performing their role, consider whether the service can be provided remotely/virtually, if a transparent barrier can be in place, or if other modifications to the service may be made to reduce physical interaction. If none of these can be implemented, staff should practice physical distancing as possible while interacting with each cohort. This includes itinerant staff who work in multiple schools.

Masks are not a replacement for physical distancing.

###### Staff Only Spaces and Gatherings

Experience from the fall of 2020 underscores the importance of COVID-19 prevention among adults in the school setting. Attention should be given to ensuring physical distancing is practiced within staff only spaces, including during break times. To support this, schools should:

- Hold meetings, in-service and professional development activities and other gatherings virtually whenever possible. If meetings cannot be held virtually:
  - Staff should practice physical distancing for face-to-face meetings, whenever possible.
  - If physical distancing is not possible, and a barrier is not present, participants should wear masks. The number of participants gathered, and the length of the gathering should be minimized as much as possible.
- Use visual cues (floor markings, posters, etc.) to promote physical distancing in common spaces (e.g. break rooms, copy rooms, etc).

WorkSafe BC guidance for offices lists measures that should be considered and implemented as applicable to the workplace for staff in office environments (both inside and outside of school buildings). This guidance is available [here](https://www.worksafebc.com/en/about-us/covid-19-updates/covid-19-returning-safe-operation/offices).

#### Other Prevention Strategies

The following strategies should be implemented wherever and whenever possible:

- Implement strategies that prevent crowding at pick-up and drop-off times.
  - Focus on entry and exit areas, and other places where people may gather or crowd.
- Stagger recess/snack, lunch and class transition times.
- Take students outside more often, for learning and break times.
  - Playgrounds can be used as normal. Ensure appropriate hand hygiene practices before and after outdoor play.
- Incorporate more individual activities or activities that encourage greater space between students and staff.
  - For elementary students, adapt group activities to minimize physical contact and reduce shared items.
  - For middle and secondary students, minimize group activities and avoid activities that require physical contact.
- Manage flow of people in common areas, including hallways, to minimize crowding and allow for ease of people passing through.

#### Visitors

Parents, caregivers, health-care providers, volunteers, and other non-staff people (e.g. visitors) entering the school should be limited to those supporting activities that are of benefit to student learning and wellbeing (e.g. teacher candidates, immunizers, meal program volunteers, etc.).

- All visitors should provide active confirmation (e.g. sign in at entry, e-mail before entry, etc.) that they have no symptoms of illness and are not required to self-isolate before entering.
- Schools should keep a list of the date, names and contact information for all visitors who enter the school.
- All visitors should wear a mask when in the school.

#### Curriculum, Programs and Activities

###### Field Trips

Schools can continue to use alternate spaces outside of school grounds (e.g. community and recreation centres, other school facilities) and to provide field trips, aligned with the guidance included in this document, relevant Provincial Health Officer Orders, and any other site-specific guidance.

Overnight or international field trips should not occur at this time.

###### Music Education

Students within the same cohort should be spaced as far apart as possible. In middle and secondary schools, masks should be worn when singing.

Music education should occur in line with guidance developed by the British Columbia Music Educators’ Association and the Coalition for Music Education in British Columbia, available [here](https://drive.google.com/file/d/1KG2rE1rU-NENxbQsuYN20xnM9TBlNn3Z/view).

###### Physical Education

People should be spread out as far as possible during physical activity. Activities should be adapted wherever possible to reduce physical contact. There should be no activities that include prolonged physical contact (i.e. physical contact beyond a brief moment) or crowding. For example, activities like tag or touch football are lower-risk, whereas activities like wrestling or partner dancing should be avoided.

Physical education and extracurricular exercise and sport activities should occur outside whenever possible.

High intensity exercise activities are those that result in significantly increased respiration rates. In middle and secondary schools:

- If indoors and the activity is stationary, have students spaced 2 metres apart. If the activity involves movement, ensure there is ample space available to reduce the likelihood of physical contact beyond a brief moment.
- Move activities outside or pursue a low-intensity activity if this isn’t possible.

Masks should be worn indoors by middle and secondary students during low-intensity indoor activities where physical distancing cannot be consistently practiced. Wearing masks during high intensity exercise activities or outdoors is based on personal choice, but cannot be in place of the other measures detailed in this section.

Masks should be worn by staff during physical education when they are unable to practice physical distancing.

Shared equipment can be used; it should be cleaned according to the Cleaning and Disinfection section of this guidance.

###### Extracurricular Activities

School-based extracurricular activities including sports, arts or special interest clubs can occur if they can be implemented in line with the guidance for within- and outside-of-cohort interactions noted in this document.

Intra-school events that are not an educational activity (i.e. are not offered specifically for student learning, health and development or mental well-being and inclusion) are considered events as defined by the Provincial Health Officer Order on [Gatherings and Events](https://www2.gov.bc.ca/assets/gov/health/about-bc-s-health-care-system/office-of-the-provincial-health-officer/covid-19/covid-19-pho-order-gatherings-events.pdf). For that reason, they must comply with this Order. This includes events like student dances, music, theatrical or dance performances, parties, services, or other occasions where large groups of people may gather and health and safety guidelines may be difficult to implement.

Inter-school events including competitions, tournaments and festivals, should not occur at this time.

#### Student Transportation on School Buses

School buses used for transporting students should be cleaned and disinfected according to the guidance provided in the BCCDC’s [Cleaning and Disinfectants for Public Settings](http://www.bccdc.ca/Health-Info-Site/Documents/CleaningDisinfecting_PublicSettings.pdf) document. Additional guidance is available from [Transport Canada](https://www2.tc.gc.ca/en/services/road/federal-guidance-school-bus-operations-during-covid-19-pandemic.html).

Bus drivers should clean their hands often, including before and after completing trips. They are encouraged to regularly use alcohol-based hand sanitizer with at least 60% alcohol during trips, as well as wear a mask when they cannot practice physical distancing or be behind a physical barrier in the course of their duties.

Students should clean their hands before they leave home to take the bus, when they leave school prior to taking the bus, and when they get home.

The following is recommended for buses:

- Open windows when the weather permits.
- If space is available, students should each have their own seat.
  - They should be seated beside the window.
- Use consistent and assigned seating arrangements.
  - Consider the order students typically onload and offload to support buses being loaded from back to front and offloaded from front to back.
  - Prioritize students sharing a seat with a member of their household or cohort.
  - The seating arrangement can be altered whenever necessary to support student health and safety (e.g. accommodating children with a physical disability, responding to behavioural issues, etc.).
- All K-12 staff and middle and secondary students should wear masks.
  - These should be put on before loading.

Additional measures can be taken, including:

- Encouraging active transportation (e.g. biking, walking, etc.) or private vehicle use by students and staff where possible to decrease transportation density.

Schools/school districts should keep up-to-date passenger lists to share with public health should contact tracing need to occur.

Other transportation methods not listed here can be used, with this guidance adapted as relevant to their mode of transportation (e.g. vans, boats, ferries, etc.).

Food Services
Schools can continue to include food as part of learning and provide food services, including for sale, if food is prepared:

- as part of learning and is consumed by the student(s) who prepared it, no additional measures beyond those articulated in this document and normal food safety practices need to be implemented (e.g. home economics and culinary arts).
- for meal programs, breakfast clubs and other food access initiatives, and is not regulated under the Food Premises Regulation, no additional measures beyond those articulated in this document and normal food safety practices need to be implemented.
  - Appendix D provides additional guidance that may be useful when offering school meal programs, breakfast clubs and other food access initiatives.

[FOODSAFE](http://www.foodsafe.ca/index.html) Level 1 covers important food safety and worker safety information including foodborne illness, receiving and storing food, preparing food, serving food, and cleaning and sanitizing. It is a helpful resource for those seeking education and training on food safety practices.

Some schools offer food services that are regulated under the [Food Premises Regulation](https://www.bclaws.ca/civix/document/id/complete/statreg/11_210_99). These are typically cafeterias, though may include some meal programs, if:

- food service is provided in schools is regulated under the Food Premises Regulation, no additional measures beyond those articulated in this document and regular requirements as outlined in the regulation need to be implemented (e.g. a FOODSAFE trained staff member, a food safety plan, etc.).
  - Additional considerations that may be relevant when providing food services in schools are detailed in the [WorkSafe BC Restaurants, cafes, pubs, and nightclubs: Protocols for returning to operation](https://www.worksafebc.com/en/about-us/covid-19-updates/covid-19-returning-safe-operation/restaurant-cafes-pubs).

For food contact surfaces, schools should ensure any sanitizers or disinfectants used are approved for use in a food service application and are appropriate for use against COVID-19. These may be different than the products noted in this document for general cleaning and disinfection. Additional information is available [here](http://www.bccdc.ca/health-info/diseases-conditions/covid-19/employers-businesses/food-businesses).

Schools can continue to accept food donations to support learning and the delivery of meal programs, breakfast clubs and other food access initiatives.

Students may be facing increased levels of food insecurity (a worry or lack of financial means to buy healthy, safe, personally acceptable food). Wherever possible, schools are encouraged to continue providing meal programs, breakfast clubs, and other food access initiatives.

The December 30, 2020 Order of the Provincial health Officer [Food and Liquor Serving Premises and Retail Establishments Which Sell Liquor](https://www2.gov.bc.ca/assets/gov/health/about-bc-s-health-care-system/office-of-the-provincial-health-officer/covid-19/covid-19-pho-order-nightclubs-food-drink.pdf) does not apply to schools. [Food Safety Legislation](https://www2.gov.bc.ca/gov/content/health/keeping-bc-healthy-safe/food-safety/food-safety-legislation) and the [Guidelines for Food and Beverage Sales in BC Schools](https://www2.gov.bc.ca/assets/gov/education/administration/kindergarten-to-grade-12/healthyschools/2015_food_guidelines.pdf) continue to apply as relevant.

Schools should emphasize that food and beverages should not be shared.

#### Community Use of Schools

Community use of school facilities must be in compliance with relevant [Orders from the Provincial Health Officer](https://www2.gov.bc.ca/gov/content/health/about-bc-s-health-care-system/office-of-the-provincial-health-officer/current-health-topics/covid-19-novel-coronavirus), including the [Events and Gathering Order](https://www2.gov.bc.ca/assets/gov/health/about-bc-s-health-care-system/office-of-the-provincial-health-officer/covid-19/covid-19-pho-order-gatherings-events.pdf) and any other related guidance.

### Personal Measures

#### Self Isolation and Illness

###### Stay Home When Required to Self-Isolate

The following students, staff or other persons **must stay home and** [**self-isolate**](http://www.bccdc.ca/health-info/diseases-conditions/covid-19/self-isolation) **as per public health direction:**

- A person confirmed by public health as a case of COVID-19; or
- A person confirmed by public health as a close contact of a confirmed case or outbreak of COVID-19; or
- A person who has travelled outside of Canada in the last 14 days.

Anyone required to self-isolate will be supported by public health. Additional information is available from [BCCDC](http://www.bccdc.ca/health-info/diseases-conditions/covid-19/self-isolation).

###### Stay Home When Sick

Students, staff, and other persons in the school should stay home when they are sick.

###### Daily Health Check

School administrators should ensure:

- Staff and other adults entering the school are aware they should not come to school if they are sick or are required to self-isolate.
- Parents and caregivers are aware that their child should not come to school if they are sick or are required to self-isolate as per public health direction.

School administrators can support this practice by communicating the requirement for everyone to do a **daily health check.**

- For school staff, an active daily health check must be completed in line with the requirements of the Provincial Health Officer’s [Order on Workplace Safety](https://www2.gov.bc.ca/assets/gov/health/about-bc-s-health-care-system/office-of-the-provincial-health-officer/covid-19/covid-19-pho-order-workplace-safety.pdf). WorkSafe BC resources to support this can be found [here](https://www.worksafebc.com/en/about-us/covid-19-updates/health-and-safety/health-checks).
  - Other adults in the school should also complete an active daily health check.
- For students, this means ensuring their parent or caregiver is aware of common symptoms of COVID-19 and is checking with their child daily to see if the child is experiencing any of these symptoms, as well as ensuring their child is not required to self-isolate.

The Ministry of Education’s [K-12 Health Check](https://www.k12dailycheck.gov.bc.ca/) app and the [When To Get Tested for COVID-19](about:blank) resource can be used to support daily health checks for students.

If the staff or student (or their parent) indicates that the symptoms are consistent with a previously diagnosed health condition and are not unusual for that individual, they can continue to attend school. No assessment or note should be required from a health care provider.

Those experiencing symptoms of illness can also use the [BC Self-Assessment Tool](https://bc.thrive.health/).

###### What To Do When Sick

Staff, students, and other persons entering the school are expected to follow the guidance from [BCCDC](http://www.bccdc.ca/health-info/diseases-conditions/covid-19/testing/testing-information). This is outlined in the [When To Get Tested for COVID-19](http://www.bccdc.ca/Health-Info-Site/Documents/COVID_public_guidance/When_to_get_tested.pdf) resource. Nobody should come to school if they are sick.

###### What To Do When Symptoms Develop At School

If a staff member, student or other person develops symptoms at school, follow the guidance in Appendix E, What To Do If A Student Or Staff Member Develops Symptoms At School.

###### Returning to School After Sickness

When a staff, student or other persons entering the school can return to school depends on the type of symptoms they experienced as outlined in the [When To Get Tested for COVID-19](http://www.bccdc.ca/Health-Info-Site/Documents/COVID_public_guidance/When_to_get_tested.pdf) resource.

If based on their symptoms a test was not recommended (i.e. the guidance is to ‘stay home until you feel better’), the person can return to school when their symptoms improve and they feel well enough.

If based on their symptoms a test is recommended (i.e. the guidance includes ‘get tested’), the person must stay home until they receive their test result.

- If the test is **negative**, they can return to school when symptoms improve and they feel well enough.
- If the test is **positive**, they must follow direction from public health on when they can return to school.

Staff, students and parents/caregivers can also use the [BC Self-Assessment Tool](https://bc.thrive.health/) app, call 8-1-1 or their health care provider for guidance.

###### Other Considerations for Managing Illness at Schools

- Establish procedures for those who become sick at school to go home as soon as possible.
  - Some students or staff may not be able to be picked up immediately. As such, consider having a space available where the student or staff member can wait comfortably, which is safe and is separated from others. This can include being in the same room as others, as long as the person experiencing illness is at least 2 metres away from others and wears a mask if they’re able to. Provide supervision for younger children.
- **Do not require a health-care provider note (i.e. a doctor’s note) to confirm the health status of any individual, beyond those required to support medical accommodation as per usual practices**.

Students or staff may still attend school if a member of their household develops new symptoms of illness, provided the student/staff has no symptoms themselves. If the household member tests positive for COVID-19, public health will advise the asymptomatic student/staff on quarantine or self-isolation and when they may return to school. Most illness experienced in BC is not COVID-19, even if the symptoms are similar.

#### Hand Hygiene

Rigorous hand washing with plain soap and water reduces the spread of illness. Everyone should practice diligent hand hygiene.

**How to practice diligent hand hygiene:**

- Wash hands with plain soap and water for at least 20 seconds. Antibacterial soap is not needed for COVID-19.
  - Temperature does not change the effectiveness of washing hands with plain soap and water, though warm water is preferred for personal comfort.
- If sinks are not available (e.g., students and staff are outdoors), use alcohol-based hand rub containing at least 60% alcohol.
  - See the [List of Hand Sanitizers Authorized by Health Canada](https://www.canada.ca/en/health-canada/services/drugs-health-products/disinfectants/covid-19/hand-sanitizer.html) for products that have met Health Canada’s requirements and are authorized for sale in Canada.
- If hands are visibly soiled, alcohol-based hand rub may not be effective at eliminating microbes. Soap and water are preferred when hands are visibly dirty. If it is not available, use an alcohol-based hand wipe followed by alcohol-based hand rub.
- To learn about how to perform hand hygiene, please refer to the BCCDC’s [hand washing poster](http://www.bccdc.ca/health-professionals/clinical-resources/covid-19-care/signage-posters).

**Strategies to ensure diligent hand hygiene:**

- Facilitate regular opportunities for staff and students to practice hand hygiene.
  - Use portable hand-washing sites or alcohol-based hand rub dispensers where sinks are not available.
- Promote the importance of diligent hand hygiene to staff and students regularly.
  - Use posters and other methods of promotion.
- Ensure hand washing supplies are well stocked at all times including soap, paper towels and where appropriate, alcohol-based hand rub with a minimum of 60% alcohol.
- Staff should assist younger students with hand hygiene as needed.

An information sheet on when students and staff should practice hand hygiene is included as Appendix F.

#### Respiratory Etiquette

Students and staff should:

- Cough or sneeze into their elbow or a tissue. Throw away used tissues and immediately perform hand hygiene.
- Refrain from touching their eyes, nose or mouth with unwashed hands.
- Refrain from sharing any food, drinks, unwashed utensils, cigarettes, or vaping devices.

Parents and staff can teach and reinforce these practices among students.

#### Water Stations and Fountains

Students and staff should be encouraged to bring an individual, filled water-bottle or other beverage container to school each day for their personal use to support hydration needs.

Re-filling water stations can be used to re-fill personal containers. These should not include bathroom sinks or other water sources not typically used for drinking water.

Water fountains where a person drinks directly from the spout should be used minimally, and only if no other means of water access are available. Hand hygiene should be practiced before and after use.

#### Personal Items and School Supplies

Students and staff can continue to bring personal items and school supplies to school for their own use. This includes reusable food containers for bringing drinks, snacks and meals.

Items brought regularly to and from school should be limited to those that can be easily cleaned (e.g. reusable food containers) and/or are considered to be low risk (e.g. clothing, paper, etc.).

### Personal Protective Equipment (PPE)

#### Non-Medical Masks and Face Coverings (Masks)

Although personal protective equipment (including masks) is low on the Hierarchy of Infection Prevention and Exposure Control Measures, it can provide an additional layer of protection when more effective measures are not feasible. Masks have a role to play in preventing the spread of COVID-19. They provide some protection to the wearer and to those around them. The term ‘mask’ in this document means a non-medical mask or face covering. Medical-grade masks are not recommended within school settings for general use.

Masks do not prevent the spread of COVID-19 on their own. They should not be used in place of physical distancing or any other measures noted in this guidance. Masks can be safely worn by school-aged children.

Based on our understanding of COVID-19 in children and adults, in schools:

Elementary students’ mask use should be based on their personal or family/caregiver’s choice.

K-12 staff and middle/secondary students should wear a mask indoors at school except when:

- Sitting or standing at their seat or workstation in a classroom or learning space,
- There is a barrier in place,
- Eating or drinking.

K-12 staff and middle/secondary students should wear a mask on buses.

Specific guidance for mask use during physical education and extracurricular exercise and sports activities is detailed here. Specific guidance for mask use during music education is detailed here. Masks don’t need to be worn outdoors.

Further guidance for staff use of masks in office settings (both within school buildings, as well as in other office settings) is available from [WorkSafe BC](https://www.worksafebc.com/en/resources/health-safety/information-sheets/covid-19-health-safety-selecting-using-masks?lang=en).

Those wearing masks must still seek to practice physical distancing. There must be no crowding or congregating of people, even if masks are worn.

Masks should not be used in place of the other measures detailed in this document.

Students should not be required to wear a mask if they do not tolerate it (for health or behavioural reasons). Schools are encouraged to support student mask use through positive and inclusive approaches, and not punitive measures or enforcement activities that exclude students from fully participating in school or that could result in stigma.

**Do not require a health-care provider note (i.e. a doctor’s note) to determine if a person does not tolerate a mask**.

Face shields are a form of eye protection for the person wearing it. They may not prevent the spread of droplets from the wearer. Face shields should not be worn in place of masks, except for those communicating using lip-reading, when visual facial cues are essential, or when people may be unable to wear a mask. Clear masks that cover the nose and mouth are another option when visual communication is necessary.

Additional information about types of masks and how to wear them is available from [BCCDC](http://www.bccdc.ca/health-info/diseases-conditions/covid-19/prevention-risks/masks).

#### PPE When Providing Student Services

###### Students with Medical Complexity, Immune Suppression and/or Receiving Delegated Care

Supporting students with medical complexities, immune suppression, or receiving delegated care may require those providing health services (e.g. staff providing delegated care or other healthcare providers) to be in close physical proximity or in physical contact with a medically complex or immune compromised student for an extended period of time. Those providing health services in schools should wear a mask (medical or non-medical) when providing services when those services cannot be provided from behind a barrier. Additional PPE over and above what is needed for routine practices is not necessary.

While implementation of infection prevention and exposure control measures help create a safe environment by helping to significantly reduce the risk of COVID-19 transmission, it does not eliminate the risk entirely. Parents and caregivers of children who are considered at higher risk of severe illness due to COVID-19 are encouraged to consult with their health- care provider to determine their child’s level of risk. Additional information is available from [BCCDC](http://www.bccdc.ca/health-info/diseases-conditions/covid-19/priority-populations).

###### Students with Disabilities and Diverse Abilities

Supporting students with disabilities and diverse abilities may require those providing services to be in close physical proximity or in physical contact with a student for an extended period of time. Those providing these services should wear a non-medical mask when providing services when the service cannot be provided from behind a barrier.

Face shields can be worn in place of masks when communicating using lip-reading, when visual facial cues are essential, or when people may be unable to wear a mask due to a medical condition. Clear masks that cover the nose and mouth are another option when visual communication is necessary.

Those providing services to students with medical complexity, immune suppression, receiving delegated care, or with disabilities and diverse abilities should follow their standard risk assessment methods to determine if additional PPE is needed, in accordance with routine practices.

No health services should be provided to a student in school who is exhibiting any symptoms of COVID-19 (beyond those detailed if a student develops symptoms at school, as detailed in Appendix E).

#### Additional PPE

Additional PPE, such as gloves and eye goggles, are not needed for most staff beyond that used as part of routine practices for the hazards normally encountered in their regular course of work.

February 4, 2021

COVID-19 Public Health Guidance for K-12 Schools

### Appendix A: Evidence Summary

###### COVID-19 in BC

- BC currently has variable community prevalence of COVID-19; some parts of the province have relatively low community transmission while other parts have relatively high levels of community transmission.
  - Since symptom-based testing began on April 21, over 1.5 million tests have been conducted in BC. While the proportion of people testing positive changed over time relative to the prevalence in the community, most people getting tested with COVID-19 like symptoms do not have COVID-19.
    - As of January 30, 2021, there were 66,779 confirmed cases in BC.
  - For those who are positive, the likely source of transmission for approximately 2 out of 3 people was a known, confirmed COVID-19 case (i.e. not community transition).

###### COVID-19 and Children

- Most children are not at high risk for COVID-19 infection.
- COVID-19 virus has a relatively low infection rate among children (ages 0 to 18). In BC, from September 7 to December 31, 2020:
  - ~3% of younger children (aged 5-12) and ~6% of older children (13-18) tested for COVID-19 were positive;
  - ~12% of all confirmed cases of COVID-19 were among children (0-18), despite this group making up ~20% of the general population.
  - Younger children (aged 5-12) comprise a smaller proportion of the total confirmed child cases compared to children between the ages of 13 and 18.
- To the end of December 2020, 47 children under 18 were admitted to hospital for COVID-19 in BC. There have been no deaths.
- Children do not appear to be the primary drivers of COVID-19 transmission in schools, community settings or households.
  - Based on published literature to date, the majority of cases in children are the result of household transmission from an asymptomatic adult family member with COVID-19. Within households and family groupings, adults appear to be the primary drivers of transmission. Older children are more likely to transmit than younger children.
- Children under one year of age, and those who are immunocompromised or have pre-existing pulmonary conditions are at a higher risk of more severe illness from COVID-19 (visit the [BCCDC Children with Immune Suppression](http://www.bccdc.ca/health-info/diseases-conditions/covid-19/priority-populations/children-with-immune-suppression) page for further details).
  - Children who are at higher risk of severe illness from COVID-19 can still receive in-person instruction. Parents and caregivers are encouraged to consult with their health-care provider to determine their child’s level of risk. Additional information is available [here](http://www.bccdc.ca/Health-Professionals-Site/Documents/COVID19-easing-social-distrancing-IS-children.pdf%20gatherings).
  - Children who have health conditions that may place them at increased risk for more severe outcomes should speak to their health care provider to determine their individual level of risk.
- Children can experience the same [symptoms](http://www.bccdc.ca/health-info/diseases-conditions/covid-19/about-covid-19/symptoms) as adults but may show symptoms differently. For example, fatigue may show in children as poor feeding, decreased activity, or changes in behaviour.

###### COVID-19 and Adults

- Most of the people infected with COVID-19 in B.C. from September 8 to December 31^st^ were adults (19 years or above). Adults represented ~88% of the cases, though make up ~82% of the population.
- Some adults with specific health conditions are at an increased chance of developing severe illness or complications from COVID-19, including older people and those with chronic health conditions. Additional information is available from [BCCDC](http://www.bccdc.ca/health-info/diseases-conditions/covid-19/priority-populations).
- Adults who have health conditions that may place them at increased risk for more severe outcomes should speak with their health care provider to determine their individual level of risk.

###### COVID-19 and Schools

- Schools do not appear to result in significant increases in community transmission of COVID-19.
  - The likelihood of a person attending school while infectious with COVID-19 reflects local community prevalence.
  - Increasing evidence supports that widespread asymptomatic transmission is not driving transmission in schools.
  - Internationally, transmission within schools accounts for a minority of all COVID-19 cases.
- Implementation of infection prevention and exposure control measures is critically important to limiting the spread of COVID-19 in schools.
  - The risk of transmission in school settings is low when infection prevention and exposure control measures are in place and adhered to.
- Internationally, in-person attendance at school in the two weeks preceding a positive test has not been associated with increased likelihood of infection, as people who tested positive were more likely to have attended social activities and gatherings with others outside of the household.
- Within BC:
  - School medical health officers note that most school exposures did not result in transmission within the school. When transmission occurred, it typically resulted in a small number of additional cases.
    - In [Vancouver Coastal Health](http://www.vch.ca/about-us/news/vch-reports-on-covid-19-transmission-in-schools-across-its-region), from September 10^th^ to December 18^th^, approximately 700 students or staff (out of a total population of over 100 thousand), were diagnosed with COVID-19. Over 90 per cent of these cases did not result in any school-based transmission.
- For adults working within schools:
  - transmission from staff to staff is more likely than among staff to students, students to staff, or students to students.
  - There does not appear to be a higher risk of COVID-19 at school than in the community or in their household.
  - There does not appear to be a higher risk of COVID-19 than other occupations that involve contact with others.
- The detection of multiple COVID-19 cases within a school does not mean that transmission occurred within the school setting; these can be related to exposure within the community and households.
- Schools and childcare facility closures have significant negative mental health and socioeconomic impacts on children, including increased stress, and decreased educational outcomes, connectedness with peers and the broader community, and health behaviours. These outcomes disproportionately impact children with vulnerabilities.
- Prevention measures and mitigation strategies involving children must be commensurate with risk.

This information is based on the best evidence currently available and will continue to be updated.

For up-to-date information on COVID-19, visit the [BC Centre for Disease Control (BCCDC) website.](http://www.bccdc.ca/health-info/diseases-conditions/covid-19)

### Appendix B: COVID-19 School Health and Safety Checklist

**Open** [COVID-19 School Health and Safety Checklist (PDF)](http://www.bccdc.ca/schools/Documents/Health_Safety_Checklist.pdf)

Complete this checklist with your school’s health and safety committee to assess your school’s safety plan with the [Provincial COVID-19 Health and Safety Guidelines for K-12 Settings](https://www2.gov.bc.ca/assets/gov/education/administration/kindergarten-to-grade-12/safe-caring-orderly/k-12-covid-19-health-safety-guidlines.pdf). Measures below should always be in place.

**SCHOOL:_________________________________________	DATE:____________________________**

| Administrative Measures | | |
| --- | --- | --- |
| **Entrance and Exits** | Staff and students are not crowded when they enter and leave the school. This includes spaces like hallways, coat rooms and bus waiting areas. | € Yes  € Sometimes  € No |
| **Common Spaces** | Staff and students can move through common spaces - hallways, washrooms, cafeteria, bus waiting areas - without crowding or physical contact. | € Yes  € Sometimes  € No |
| **Physical Distancing Within Learning Groups** | Physical contact and close, face-to-face interactions are minimized. People are spread out as much as possible. | € Yes  € Sometimes  € No |
| **Physical Distancing Outside of Learning Groups** | There is 2 meters of space available between people from different learning groups when together for extended periods of time (when indoors for elementary, and at all times for middle and secondary). | € Yes  € Sometimes  € No |
| **Staff Only Common Spaces (e.g. Break Rooms, Copy Rooms, etc.).** | Physical distancing is practiced. Masks are not used in place of physical distancing. | € Yes  € Sometimes  € No |
|  | Visual cues (floor markings, posters, etc.) are in place to promote physical distancing. | € Yes  € Sometimes  € No |
|  | Masks are worn in accordance with the Health and Safety Guidelines. | € Yes  € Sometimes  € No |
| **Itinerant Staff, Temporary Teachers on Call and Other Visitors** | Process in place to ensure itinerant staff, Teachers On-Call and visitors are aware of the school’s health and safety measures and their responsibility to follow them at all times. | € Yes  € Sometimes  € No |
| **Gatherings** | Student gatherings (e.g. events that bring staff and students together outside of regular learning activities) only occur within learning groups and as minimally as possible. | € Yes  € Sometimes  € No |
|  | Staff gatherings (e.g. meetings, professional development activities, etc.) occur virtually whenever possible. If not possible, staff are able to be physically distanced during the meeting. | € Yes  € Sometimes  € No |
| **Extracurricular Activities** | Activities are implemented in line with the guidance for within- and outside-of-learning group interactions, including 2 meters of space available between people from different learning groups (when indoors for elementary, and at all times for middle and secondary). | € Yes  € Sometimes  € No |
| **Hand Hygiene** | Hand cleaning facilities are available and accessible throughout the school and are well maintained. | € Yes  € Sometimes  € No |
|  | Signage to remind students staff to practice regular hand hygiene and good cough etiquette. | € Yes  € Sometimes  € No |

| Environmental Measures | | |
| --- | --- | --- |
| **Learning Space Configuration** | Learning spaces are arranged to maximize the space available and to minimize people directly facing one another (where possible). | € Yes  € Sometimes  € No |
| **Increased Cleaning and Disinfecting** | General cleaning and disinfecting is done every 24 hours, with frequently-touched surfaces cleaned an additional time (including once during the school day). | € Yes  € Sometimes  € No |
| **Ventilation and Air Exchange** | The school’s ventilation system is serviced and operating to specifications. | € Yes  € Sometimes  € No |

| Personal Measures | | | | |
| --- | --- | --- | --- | --- |
| **Daily Health Checks** | | Staff complete an active Daily Health Check. | | € Yes  € Sometimes  € No |
|  |  | Parents and students are reminded of their responsibilities to complete a Daily Health Check. | | € Yes  € Sometimes  € No |
| **Stay Home When Sick** | | Staff and students are reminded to stay home when they are sick. | | € Yes  € Sometimes  € No |
| Personal Protective Equipment | | | | |
| **Masks** | Staff, itinerant staff, and visitors (who are able to), wear masks in accordance with the K-12 Health and Safety Guidelines. | | € Yes  € Sometimes  € No | |
|  | Students (who are able to), wear masks in accordance with the Health and Safety Guidelines. | | € Yes  € Sometimes  € No | |
|  | Masks are available for those who have forgotten theirs. | | € Yes  € Sometimes  € No | |

### Appendix C: Contact Tracing and Public Health Notifications in Schools

Student or staff confirmed to be COVID-19 Case

Confirmed case interviewed by public health to determine how they were infected and who they were in close contact with while infectious

If confirmed case was not infectious when they attended school, the school will not be contacted.

If confirmed case was infectious while they attended school OR it's determined that they were infected at school, public health will inform a school/district administrator and:

May request class, cohort or bus lists

Determine if anyone at school was potentially exposed

Contact those potentially exposued directly with letter or phone call

Confirmed close contacts asked to self-isolate

At health authority discretion: Non-close contacts asked to monitor for symptoms

Coordinate with the school/district to support communications

Exposure notification posted on health authority websites

At health authority discretion: provide exposure notification letter to the broader school community

Regional health authorities determine their own notification processes. The notifications used in some regions may differ from what is included here. In all regions, public health ensures anyone who may be a close contact (i.e. those required to self-isolate) is notified. Additional information on contact tracing, self-isolation and close contacts is available from [BCCDC](http://www.bccdc.ca/health-info/diseases-conditions/covid-19).

### Appendix D: Supplementary Guidance for School Meal Programs

This guidance is adapted from the [WorkSafe BC Restaurants, cafes, pubs, and nightclubs: Protocols for returning to operation](https://www.worksafebc.com/en/about-us/covid-19-updates/covid-19-returning-safe-operation/restaurant-cafes-pubs) to support the delivery of school meal programs, breakfast clubs and other food access initiatives that are not regulated under the *Food Premises Regulation*.

###### General Considerations

- Students from different cohorts can access school meal programs at the same time if necessary (e.g. a morning breakfast program offered only to students who may need it). Physical distance between students from different cohorts should be maintained as much as is practical to do so while ensuring the program can be offered.

###### Food Delivery and Preparation

- Limit the number of staff/volunteers in a food preparation or eating area at any one time to those necessary to ensure the program can be delivered.
- Inform delivery agents and other volunteers of how to adhere to the school’s visitor policy, where food should be delivered to, and what hours food can be accepted at.
- Develop and establish hand hygiene procedures for all staff/volunteers. This includes before and after leaving the food preparation area and using equipment.
- Donated food, including Traditional foods, can continue to be accepted in line with regular food safety precautions for accepting food donations.

###### Cleaning and Disinfecting

- Continue with regular cleaning and disinfecting practices for food services.
- Identify high-touch surfaces to ensure they are cleaned and disinfected in line with the guidance in this document and existing food safety practices.
  - High-touch surfaces may include ingredients and containers, equipment such as switches, dials and handles and shared serving utensils if they are used by multiple people.

###### Food Distribution to Students

- Students should practice hand hygiene before accessing food.
- Schools can continue to provide self-service stations (e.g., salad bar, self-serve breakfast, etc.).
  - Consider pre-plating or serving food directly if students are unable to consistently implement personal measures (e.g. practice regular hand hygiene, not touch their face, etc.) or to prevent gathering or crowding.
- Post signs to remind students to practice hand hygiene and to maintain space from one another.
- If food is served to students, re-usable plates, utensils and containers can be used, with normal cleaning and disinfecting methods for dishwashing implemented.
- Provided food safety precautions are followed, leftover food can be sent home with students.

### Appendix E: What to Do if a Student or Staff Member Develops Symptoms At School

| **If a Student Develops Symptoms of Illness At School** | **If a Staff Member Develops Symptoms of Illness At School** |
| --- | --- |
| **Staff must take the following steps:**   1. Immediately separate the symptomatic student from others in a supervised area. 2. Contact the student’s parent or caregiver to pick them up as soon as possible. 3. Where possible, maintain a 2-metre distance from the ill student. If not possible, staff should wear a mask if available and tolerated, or use a tissue to cover their nose and mouth. 4. Provide the student with a mask or tissues to cover their coughs or sneezes. Throw away used tissues as soon as possible and perform hand hygiene. 5. Avoid touching the student’s body fluids (e.g., mucous, saliva). If you do, practice diligent hand hygiene. 6. Once the student is picked up, practice diligent hand hygiene. 7. Staff responsible for facility cleaning must clean and disinfect the space where the student was separated and any areas recently used by them (e.g., classroom, bathroom, common areas).   Parents or caregivers must pick up their child as soon as possible if they are notified their child is ill. | **Staff should go home as soon as possible.**  If unable to leave immediately:   1. Symptomatic staff should separate themselves into an area away from others. 2. Maintain a distance of 2 metres from others. 3. Use a tissue or mask to cover their nose and mouth while they wait to be picked up. 4. Staff responsible for facility cleaning must clean and disinfect the space where the staff member was separated and any areas used by them (e.g., classroom, bathroom, common areas). |
| **Students and staff should return to school according to the guidance under the** **Returning to School After Sickness** **sections of this document.**  **A health-care provider note should not be required for students or staff to return.** | |

### Appendix F: When to Perform Hand Hygiene at School

| **When Students Should Perform Hand Hygiene:** | **When Staff Should Perform Hand Hygiene:** |
| --- | --- |
| - When they arrive at school. - Before and after any breaks (e.g., recess, lunch). - Before and after eating and drinking (excluding drinks kept at a student’s desk or locker). - Before and after using an indoor learning space used by multiple cohorts (e.g. the gym, music room, science lab, etc.). - After using the toilet. - After sneezing or coughing into hands. - Whenever hands are visibly dirty. | - When they arrive at school. - Before and after any breaks (e.g. recess, lunch). - Before and after eating and drinking. - Before and after handling food or assisting students with eating. - Before and after giving medication to a student or self. - After using the toilet. - After contact with body fluids (i.e., runny noses, spit, vomit, blood). - After cleaning tasks. - After removing gloves. - After handling garbage. - Whenever hands are visibly dirty. |
