## Appendix 1b for "Symptomatic and asymptomatic transmission of SARS-CoV-2 in K-12 schools, British Columbia, April to June 2021"

### Addendum – Public Health Guidance for K-12 Schools

March 30, 2021

Due to the recent rise in cases of COVID-19 in BC, the Provincial Health Officer has recommended the following time-limited changes to the Non-Medical Masks and Face Coverings (Masks) section, and any other sections where mask guidance is provided, to the [Public Health Guidance for K-12 Schools](http://www.bccdc.ca/Health-Info-Site/Documents/COVID_public_guidance/Guidance-k-12-schools.pdf).

Effective immediately, all staff, adult volunteers and visitors, and all Grade 4 to 12 students should wear a non-medical mask or face covering (a “mask”) at all times while indoors at school, subject to the exceptions noted below.

Exceptions

The recommendations above should not apply as follows:

- To a person who is unable to wear a mask because they do not tolerate it (for health or behavioural reasons);
- To a person unable to put on or remove a mask without the assistance of another person;
- If the mask is removed temporarily for the purposes of identifying the person wearing it;
- If the mask is removed temporarily to engage in an educational activity that cannot be performed while wearing a mask (e.g. actively playing a wind instrument, high-intensity physical activity, etc.);
- If a person is eating or drinking;
- If a person is behind a barrier;
- While providing a service to a person with a disability or diverse ability (including but not limited to a hearing impairment), where visual cues, facial expressions and/or lip reading/movements are important.

Staff, adult volunteers, and all Grade 4 to 12 students should wear a mask at all times while on a bus, subject to the exceptions noted below.

Exceptions:

The recommendation above should not apply as follows:

- To a bus driver while driving;
- To a person who is unable to wear a mask because they do not tolerate it (for health or behavioural reasons);
- To a person unable to put on or remove a mask without the assistance of another person;
- If the mask is removed temporarily for the purposes of identifying the person wearing it; or
- While eating or drinking.

Kindergarten to Grade 3 students are encouraged to wear a mask at school and on buses, but should not be required to do so.

If an activity cannot be implemented in line with this guidance, it should be adapted or another activity should be selected.

Schools continue to be encouraged to support student mask use through positive and inclusive approaches, and not punitive or enforcement activities that exclude students from fully participating in school or that could result in stigma.

No student should be prevented from attending or fully participating in school if they are not wearing a mask.

This guidance has been issued along with other [extensive public health measures](https://www2.gov.bc.ca/gov/content/covid-19/info/restrictions) announced March 29, 2021, by the Provincial Health Officer. This addendum is in place until April 19, 2021, at which point it will be reviewed. Unless revised, the guidance in this addendum should be used in place of masking guidance in the [Public Health Guidance for K-12 Schools](http://www.bccdc.ca/Health-Info-Site/Documents/COVID_public_guidance/Guidance-k-12-schools.pdf).
